# Metabolic signature of the pathogenic 22q11.2 deletion identifies carriers and provides insight into systemic dysregulation

**DOI:** 10.1101/2023.01.13.23284505

**Authors:** Julie Courraud, Francesco Russo, Gonçalo Espregueira Themudo, Susan Svane Laursen, David M. Hougaard, Arieh S. Cohen, Thomas Werge, Madeleine Ernst

## Abstract

**Background:** Large deletions at chromosome 22q11.2 are known to cause severe clinical conditions collectively known as 22q11.2 deletion syndrome. Notwithstanding the pathogenicity of these deletions, affected individuals are typically diagnosed in late childhood or early adolescence, and little is known of the molecular signaling cascades and biological consequences immediately downstream of the deleted genes.

**Methods:** Here, we used targeted metabolomics to compare neonatal dried blood spot samples from individuals clinically identified as carriers of a 3MB deletion at the chromosome 22q11.2 with unaffected individuals. A total of 173 metabolites were successfully identified and used to inform on systemic dysregulation caused by the genomic lesion and to discriminate carriers from non-carriers.

**Results:** We found 84 metabolites to be differentially abundant between carriers and non-carriers of the 22q11.2 deletion. A predictive model based on all 173 metabolites achieved high Accuracy (89%), Area Under the Curve (93%), F1 (88%), Positive Predictive Value (94%) and Negative Predictive Value (84%) with tyrosine and proline having the highest individual contributions to the model as well as the highest interaction strength.

**Conclusions:** Targeted metabolomics provides insight into the molecular consequences possibly contributing to the pathology underlying the clinical manifestations of the 22q11 deletion and is an easily applicable approach to first-pass screening for carrier status of the 22q11 to prompt subsequent verification of the genomic diagnosis.

## INTRODUCTION

22q11.2 deletion syndrome (22q11.2DS) was first reported in 1965 by Dr Angelo DiGeorge, and correspondingly named DiGeorge syndrome^1^. It is the most common chromosomal microdeletion syndrome with an estimated incidence of 1 over 3,672 live births^2^. Deletions of various sizes are gathered under the 22q11.2DS name, but the most common is a 3-Mb deletion in between two low copy repeats (LCRs) zones (LCR22A and LCR22D).

Irrespective of deletion size, 22q11.2DS comes with heterogeneous clinical presentation including several very severe conditions^3^. None of the clinical presentations are specific to 22q11.2DS, and far from all carriers are clinically ascertained. In fact, a considerable proportion of 22q11.2DS (~10%) is inherited, typically from clinically un-affected or non-recognized parents carrying the deletion^3^, and has been reported to be more severely affected than *de-novo* cases. Among the clinical presentations detected in utero or at birth, congenital heart defects are the most frequent, followed by velopharyngeal insufficiency, cleft palate, or dysmorphic craniofacial features. Many disorders will only be detectable or develop later in life, such as hypocalcemia due to primary hypoparathyroidism^4^ as well as developmental disabilities, mental disorders^5,6^, and life threatening severe chronic immune deficiency^7^. While the syndrome is not curable, many of the clinical manifestations can be improved, if treated timely.

On average, 22q11.2DS is diagnosed at 9-13 years, and children with 22q11.2DS often go through a diagnostic odyssey before receiving diagnosis and appropriate care^3,6^. In fact, due to the heterogenous clinical presentation of 22q11.2DS, patients meet on average seven experts, including psychiatrists, pediatricians, surgeons, general practitioners, psychologists, and clinical geneticists before being oriented towards molecular diagnostics^8^.

As diagnosis of 22q11.2DS can only be confirmed genetically, developing techniques for early clinical diagnosis of 22q11.2DS that can be integrated into existing screening program is essential. While it has been suggested that 22q11.2DS could be detected during neonatal metabolic screening, which is standard in most countries^9^, such assays are not yet available. Consequently, finding biomarkers specific to this syndrome that are reliably detectable in neonatal dried blood spots (DBS) could greatly help the diagnosis process, leading to earlier appropriate care. In fact, *PRODH*, the gene encoding for proline dehydrogenase and among the genes deleted in 22q11.2DS, has been reported to result in hyperprolinaemia^10–12^, suggesting that patient with hyperprolinaemia should be screened for 22q11.2DS^11^.

In this work we investigate for the first time a large cohort of individuals with the 22q11.2 deletion using targeted metabolomics of neonatal DBS to inform on the molecular consequence of a large genomic lesion and to probe metabolomics screening as a diagnostic tool for the early identification of individuals carrying the 22q11.2 deletion.

## MATERIALS AND METHODS

### Materials

Methanol (MeOH), acetonitrile (ACN), isopropanol (IPA), water (H2O) and formic acid (FA) were of Optima™ LCMS-grade and were purchased from Thermo Fisher Scientific (Waltham, MA, USA).

### Study cohort

We acquired metabolic profiles of residual dried blood spots (DBS) from 406 children born between 1983 and 2012 (median 2000) collected at a median age of 5 days after birth (range [2-40] days) and stored at the Danish National Biobank at −20°C. Cases were identified as children later diagnosed with 22q11.2DS (102 females, 101 males). Controls were typically developing children matched based on sex and date of birth. An overview of the study cohort is shown in Table 1.

**Table 1.** Characteristics of study cohort.

|  | Cases (n=203) |  |  | Controls (n=203) |  |  |
| --- | --- | --- | --- | --- | --- | --- |
|  | median | range | NA | median | range | NA |
| Age at sampling (days) | 5 | [2-40] | 19 | 5 | [2-30] | 19 |
| Birth weight (g) | 3130 | [1200-5550] | 9 | 3400 | [830-5115] | 7 |
| Gestational age (weeks) | 39 | [32-42] | 106 | 39 | [33-42] | 96 |
| Month of birth* | 7 | [1-12] | 0 | 7 | [1-12] | 0 |
| Age of mothers at birth (years) | 29.6 | [18.0-42.4] |  | 29.5 | [16.6-41.3] |  |

NA = Not available data, \* January = 1 and December = 12

### Data acquisition

We measured absolute concentrations of a total of 408 compounds in the DBS using the AbsoluteIDQ p400 kit (Biocrates Life Science Ag., Innsbruck, Austria) following the manufacturer’s instructions. Metabolites from DBS were extracted according to the standard operation procedure specific for DBS provided by Biocrates. All instrumentation and preprocessing software were from Thermo Scientific (Waltham, MA, USA). Our liquid chromatography tandem mass spectrometry (LC-MS/MS) platform consisted of an autosampler CTC Combi PAL HTS TMO (two injectors), a liquid chromatography (LC) system (LX-2 with two Ultimate 3000 Dionex RS pumps and a Transcend II Valve Interface Module) and a Q-Exactive Orbitrap mass spectrometer with a HESI-II probe. Case and control sample pairs were randomized into six 96-well analytical plates, and data was acquired during May to June 2018 with no more than 2-5 days between each plate. Each sample was injected four times as instructed: two times for LC-MS and two times for flow injection analysis (FIA) data acquisition. LC-MS and FIA-based data were acquired on a dedicated injector and pump to limit technical variation. Each plate included a solvent blank, three paper blanks (blank filter paper on which dried blood spots are collected), a 7-point calibration curve (injected for LC-MS data acquisition only), and quality control (QC) samples provided by Biocrates (three concentration levels QC1-3). QC1 and QC3 were injected once, while QC2 was injected five times across the plate among the experimental samples. Four case-control pairs had to be replicated on the last plate as technical difficulties occurred during extraction; we therefore kept only the results from the second analysis on plate 6.

### Quality control and data processing

LC-MS-based data were preprocessed as instructed by Biocrates using Xcalibur (v4.1.31.9) for peak integration and after thorough optimization and manual check. The exported feature table was then imported into MetIDQ Carbon-2793 (Biocrates) along with FIA-based raw data for plate validation and concentration calculation. Concentrations were normalized based on QC2 target values in MetIDQ to reduce batch effect. All measurements were then exported along with their individual status computed by MetIDQ (e.g. “Valid”, etc.). For further quality control and preprocessing we used the MeTaQuaC R package v0.1.32.9001^13^. All measurements with a MetIDQ status other than ‘Valid’ were replaced by missing values and only compounds detected in at least 30% of the Biocrates’ QC level 2 samples were kept in the analysis. Furthermore, compounds with 100% missing values in experimental samples and compounds, which had >80% missing values in both case and control samples were removed. Finally, we batch normalised absolute concentrations through centering by subtracting the column means (omitting NAs) of each batch and scaling by the standard deviation. The final data table consisted of 173 compounds (33 out of 42 and 140 out of 366 for the LC and FIA injections respectively) and 406 samples (203 cases and controls respectively).

### Statistical analyses

To initially contrast quantities of the analyzed metabolites between 22q11.2 carriers and controls, we performed a differential abundance analysis by applying a paired Wilcoxon signed-rank test using the R function Wilcox.test from the stats R package^14^. Subsequently, we used the function p.adjust (from the stats R package) to calculate the False Discovery Rate (FDR) adjusted p-values. Metabolites having FDR-adjusted p-value < 0.05 were considered differentially abundant.

Predictive models and feature importance were built by training an eXtreme Gradient Boosting (XGBoost) model using the R packages xgboost (version 1.4.1.1)^16^ and caret (version 6.0-88)^17^. We divided the dataset in train and test sets (80% train, 20% test). Firstly, we performed an analysis including all the 173 metabolites, without tuning and using the following recommended parameters: nrounds = 500, max_depth = 6, colsample_bytree = 1, eta = 0.3, subsample = 1, gamma = 0, min_child_weight = 1. Secondly, we improved the performance of the model with all the 173 metabolites by applying a tenfold cross-validation. The final model fitted the following tuned parameters on the full training set: nrounds = 1400, max_depth = 4, eta = 0.025, gamma = 0.5, colsample_bytree = 0.8, min_child_weight = 1, subsample = 0.5. Prediction on the test set and evaluation of performance were performed and the R package pROC (version 1.18.0)^18^ was used for calculating the area under the curve (AUC) and plotting the receiver operating characteristic (ROC) curve. In addition to AUC and accuracy we provide F1, positive and negative predictive values calculated using the caret function confusionMatrix.

To discover which metabolites contributed most to the prediction of 22q11.2DS, we computed feature importance using the caret function varImp. To further validate feature importance, we performed additional analyses. We measured how important each feature was for the predictions by applying the R function FeatureImp (iml package, version 0.10.1^19^), which implements the method described by Fisher and coworkers^20^. The method works by shuffling each feature and measuring how much the performance drops according to the prediction loss/error (in our case the classification error). The prediction error is measured before and after shuffling the values of the feature and the larger the increase of the error is, the more important the feature. This process was repeated 1000 times (n.repetitions parameter set to 1000), since the higher the number of repetitions the more stable and accurate the results become.

We then explored how strongly features interact with each other in the prediction model by applying the R function Interaction from the iml package, which measures interactions through Friedman’s H-statistic^21^. The interaction measure shows how much of the variance of each feature is explained by the interaction, with values ranging from 0 (no interaction) to 1.

To evaluate the predictive value of the most important selected metabolites, we compared the results of four XGBoost models without tuning the models’ parameters (using default/recommended settings for all models). The first model included the top two most important metabolites (tyrosine and proline), the second model only the first most important metabolite (tyrosine) and the third model only the second most important metabolite (proline).

All statistical analyses were performed in R^14^ (v4.1.1) and scripts and Jupyter notebooks are publicly available at: https://github.com/SSI-Metabolomics/22q11_SupplementaryMaterial/.

## RESULTS

To investigate the potential of targeted metabolomics as a diagnostic tool for 22q11.2DS in neonatal screening, we performed a broad metabolic profiling using a validated commercial kit (see Methods for details) of neonatal blood spots from 203 individuals identified as carriers of the 3Mb deletion at chr. 22q11.2 and 203 age- and sex-matched individuals without the deletion. After quality control, data on 173 compounds were obtained and used for subsequent analyses (see Methods for details).

To initially contrast quantities of the analyzed metabolites between 22q11.2 carriers and controls, we performed a differential abundance analysis of the 173 metabolites that passed QC (see Methods for details) by applying a paired Wilcoxon signed-rank test. In total, 84 metabolites were differentially abundant (FDR-adjusted p-value < 0.05) and among them tyrosine was the most significant (FDR-adjusted p-value = 7.53e-15; Figure 1) with a highly significant lower abundance in cases versus controls. Also, proline showed a significant difference in abundance (FDR-adjusted p-value = 2.25e-03) with a slightly higher abundance in cases compared to controls. The full list of significant differentially abundant metabolites can be found in Supplementary Data 1.

**Figure 1.**
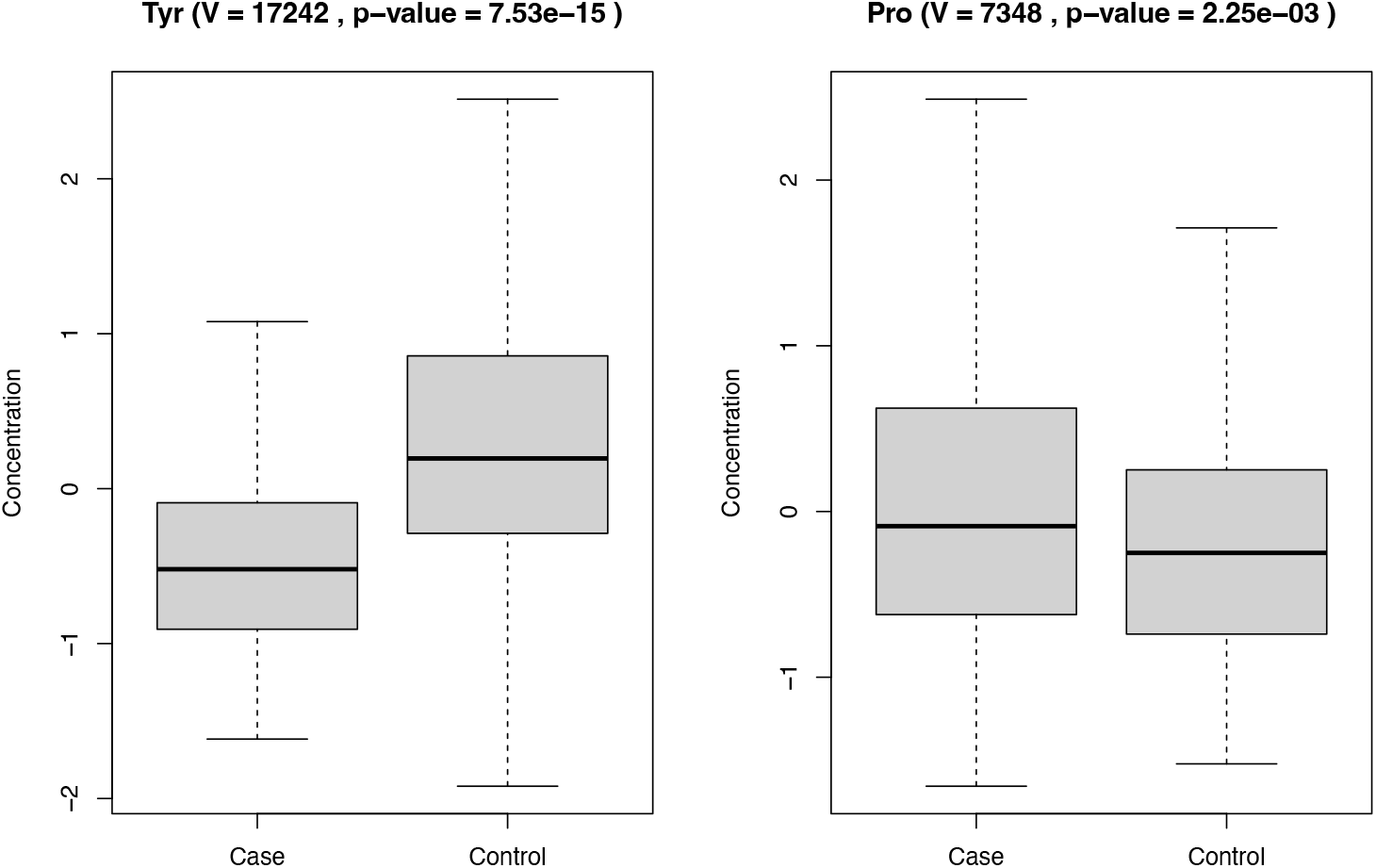
Differential abundance of tyrosine and proline. Using the Wilcoxon signed-rank test showing the comparison of cases versus controls for tyrosine (left) and proline (right). Box plots including V-statistics (V value) and FDR-adjusted p-value.

To further investigate the metabolic signature of individuals carrying the 22q11.2 deletion, we first determined the combined ability of the 173 analyzed metabolites to classifying the 406 22q11.2 case-control individuals in a predictive model, and next determined the individual contributions of each metabolite to the resulting model. As shown in Figure 2 and Table 2, the model achieved high Accuracy (89%), AUC (93%), F1 (88%), Positive Predictive Value (94%) and Negative Predictive Value (84%). The individual contributions to the final model of the top-20 metabolites are shown in Figure 3, with tyrosine and proline ranked as first and second. This finding was confirmed in a subsequent analysis, estimating the loss of performance, i.e., the classification error, as a function of shuffling measurements between metabolites^15^.

**Table 2.** Performance comparison across models. We built five models with: 1) all 173 metabolites, 2) only tyrosine and proline, 3) only tyrosine, 4) only proline. Additionally, we show the results of the tuned parameters (ten fold CV).

|  | All metabolites | Tyrosine and Proline | Tyrosine | Proline | All metabolites |
| --- | --- | --- | --- | --- | --- |
|  | No tuning | No tuning | No tuning | No tuning | Parameter tuning |
| Positive Predictive Value | 0.865 | 0.707 | 0.727 | 0.429 | 0.943 |
| Negative Predictive Value | 0.814 | 0.718 | 0.66 | 0.444 | 0.844 |
| F1 | 0.831 | 0.716 | 0.657 | 0.400 | 0.853 |
| Accuracy | 0.837 | 0.712 | 0.687 | 0.437 | 0.887 |
| AUC | 0.897 | 0.753 | 0.732 | 0.509 | 0.927 |

**Figure 2.**
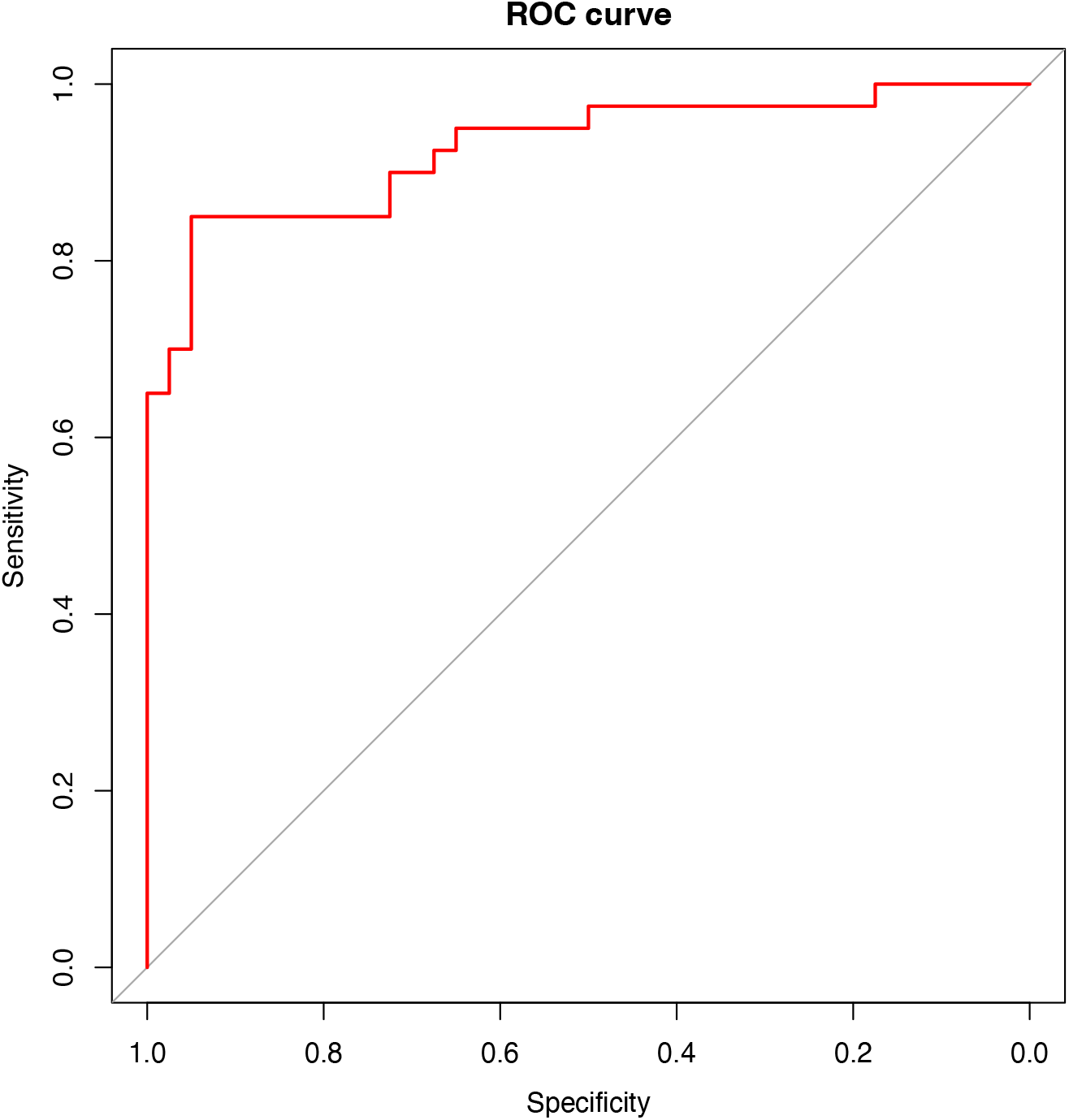
ROC curve for the predictive model. Receiver Operating Characteristic (ROC) curve showing the performance of the final classification model, including a total of 173 metabolites (ten fold CV).

**Figure 3.**
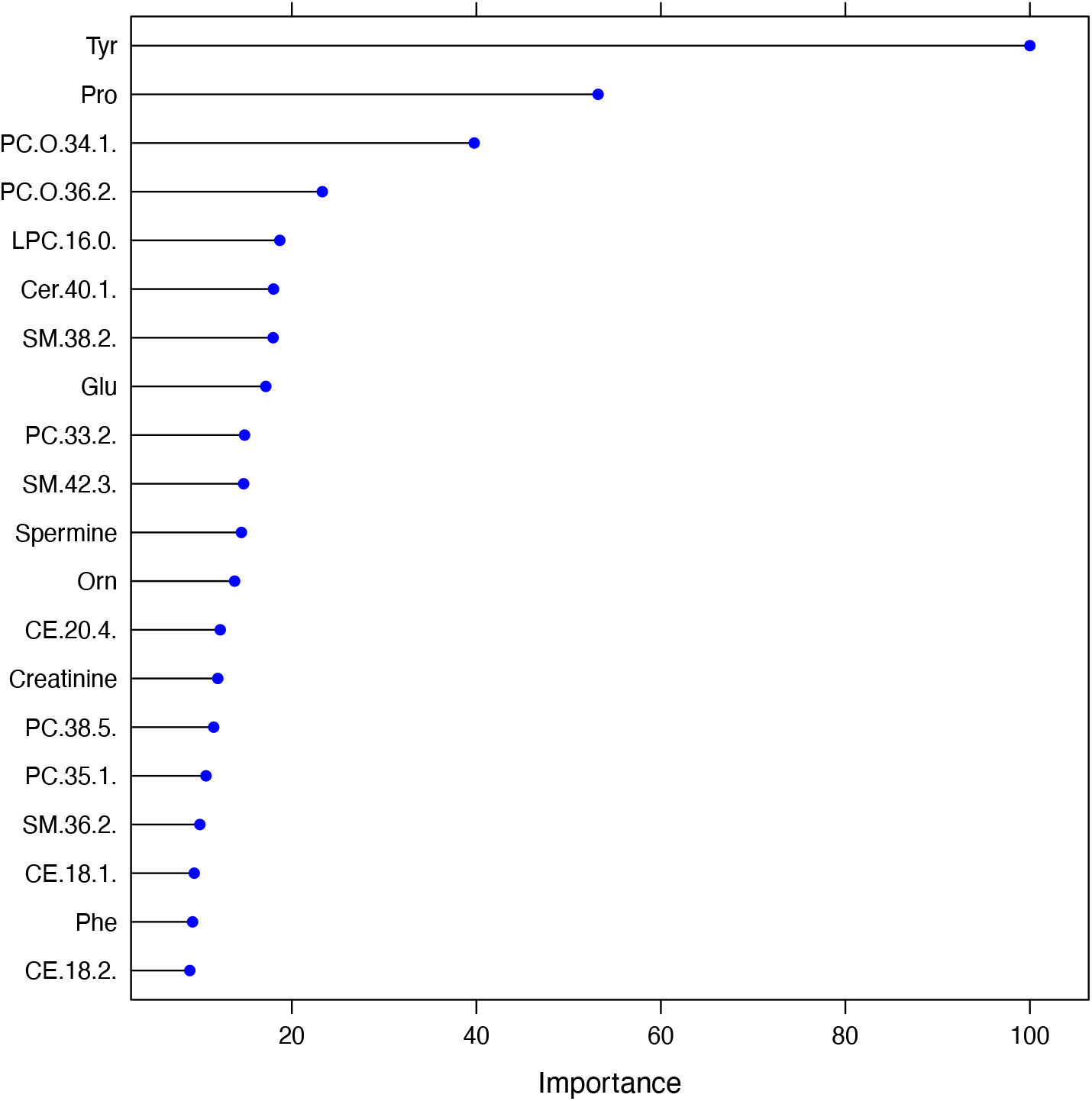
Top-20 metabolites ranked by their importance. The individual contributions to the final model of the top-20 metabolites based on the caret R function varImp, reporting a scaled measure of importance. The measure is equivalent to the Gain value (the improvement in accuracy brought by a feature to the branches it is on). The higher the value is, the more important the feature is.

Next, we examined overall interactions between the analyzed metabolites, and found tyrosine and proline to be the two metabolites with the highest interaction with other metabolites based on Friedman’s H-statistic^21^. Furthermore, tyrosine and proline were among the strongest interactors of each other, as shown in the interaction network of proline (Figure 4).

**Figure 4.**
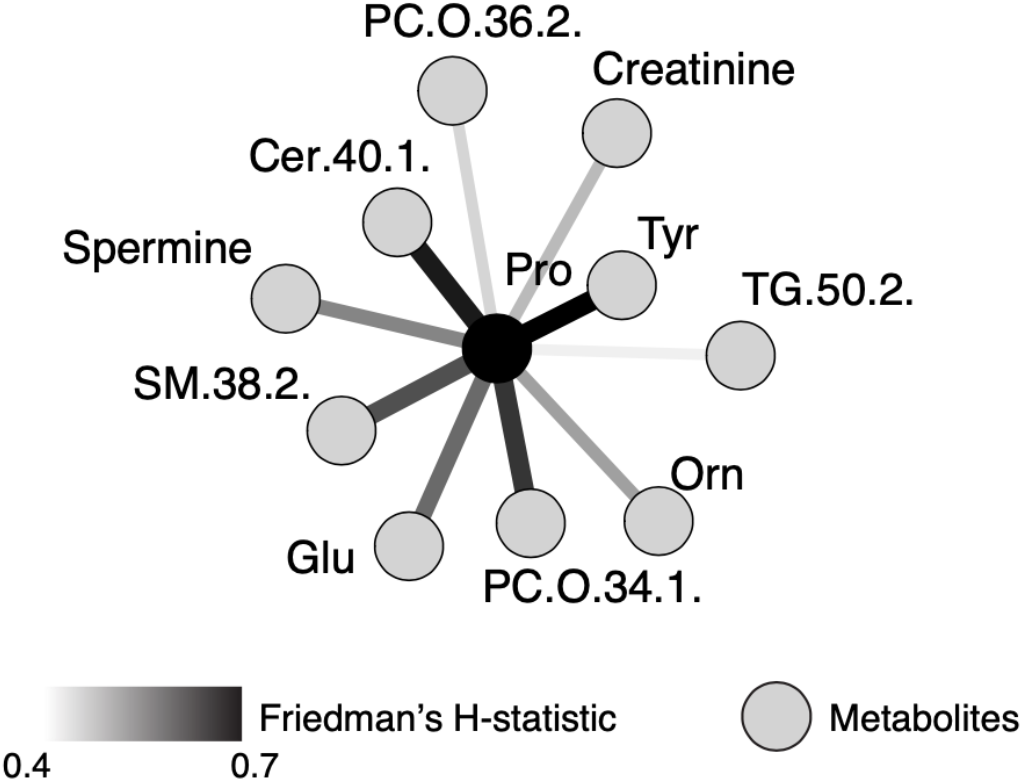
Pairwise interactions for proline. Top ten metabolites interacting most strongly with proline are shown. Edges are colored according to Friedman’s H-statistic with the strongest interaction in black and weakest interaction in light grey. The black node indicates proline, while the grey nodes are its interactors. Tyrosine shows the strongest interaction.

To contrast the individual and combined predictive values of proline and tyrosine with all the other metabolites combined, we compared the results of four XGBoost models (see Methods for details), considering (1) all 173 metabolites, (2) proline and tyrosine, (3) only tyrosine, and (4) only proline. As detailed in Table 2, these analyses show that (i) tyrosine outperforms proline on all five metrics relative to prediction of 22q11.2 status, (ii) combining tyrosine and proline improves prediction somewhat relative to tyrosine alone, while (iii) all metabolites combined increases prediction even further (Table 2).

## DISCUSSION

In this work, we explored the potential of targeted metabolomics to inform on molecular consequences of the pathogenic deletion at chromosome 22q11.2 and to serve as a neonatal diagnostic screening tool to identify individuals carrying the copy number variant. We found that 22q11.2 deletion associated with significant alterations in levels of metabolites in whole blood at time of birth, which may reflect metabolic alterations in other organ systems such as the brain. Furthermore, we used machine learning techniques and differential abundance analysis to document that the metabolite profiles discriminate 22q11.2 deletion carriers from non-carriers and identify the amino acids tyrosine and proline as the most dysregulated metabolites.

The increased levels of proline, observed in this study, is consistent with decreased proline-to-glutamate conversion due to hemizygosity of proline dehydrogenase 1 (*PRODH*) in 22q11.2 deletion carriers, and generalizes case-reports on individuals with the 22q11.2 deletion^10–12,22^. In contrast, the observation of reduced levels of tyrosine is novel and may derive from feedback inhibition of phenylalanine hydroxylase mediated conversion of dietary phenylalanine into tyrosine. Such feedback inhibition could be due to accumulating levels of catecholamines resulting from hemizygosity of the catechol-O-methyltransferase encoding *COMT*-gene in 22q11.2 deletion carriers^23^.

As demonstrated in this study, the capacity of targeted metabolomics to discriminate between individuals carrying the 22q11.2 deletion from non-carriers points to great potential for the clinical applicability of this approach in neonatal screening, which would ensure early identification and corresponding timely and proper healthcare provision and socio-educational support. Recent findings based on population-representative studies in Denmark documented that a considerable fraction of 22q11.2 deletion carriers go unnoticed in the healthcare system, even when this is public and egalitarian^2^, a shortcoming that also affects a significant proportion of carriers with other pathogenic copy-number-variants^24^.

The potential benefits and shortcomings of neonatal screening for 22q11.2 deletions have been considered in the past primarily in relation to prenatal genetic testing^25^. Advantages include early intervention to cardiac defects, hypocalcemia-induced seizures, and severe immune deficiency, while concern of causing vulnerable child syndrome through national 22q11.2 screening programs has been raised given the less-than full penetrance of the genomic aberration^25^. Importantly, these concerns contrast with ambitions to eliminate the ‘diagnostic odyssey’ and intervene against failure to thrive and early, prodromal manifestations of mental disorders; complications that are often not considered when deciding on clinical screening programs.

The strength of this study stem from the nationwide biobank from where the samples were obtained that is both representative of the population as a whole and rests on decades of national, neonatal screening for severe congenital disorders originally prompted by analyses for phenylketonuria. Importantly, the neonatal blood spots used in our study conveniently provide genomic DNA for efficient follow-up genotype-or sequence-based verification to eliminate false positive findings, which in the case of national routine screening will be a concern. In light of the ease of targeted genomic follow-up analyses, future refinement of the metabolic analyses to reduce false negative findings should be expected.

In conclusion, we identified metabolites dysregulated in 22q11.2, in particular tyrosine and proline, and document that careful metabolic profiling leveraging existing clinical screening programs could allow for the identification of individuals carrying the 22q11.2 deletion, although we emphasize that clinical performance and applicability awaits further studies.

## Data Availability

The data underlying this study are not publicly available due to the Danish Data Protection Act and European Regulation 2016/679 of the European Parliament and of the Council (GDPR) that prohibit distribution of personal data. The data are available from the corresponding authors upon reasonable request and under a data transfer and collaboration agreement.

https://github.com/SSI-Metabolomics/22q11_SupplementaryMaterial/

## ACKNOWLEDGEMENT

The iPSYCH Initiative is funded by the Lundbeck Foundation (Grant Nos. R268-2016-3925, R102-A9118, and R155-2014-1724) and approved by the Danish Scientific Ethical Committees. This research has been conducted using the Danish National Biobank resource supported by the Novo Nordisk Foundation.

## ETHICAL APPROVAL

The scientific board of the Danish Cytogenetic Central Register approved the study and provided access to the genetic data, that provided information on individuals with a 22q11.2 deletion and was obtained from the Danish Cytogenetic Central Registry (DCCR), which was established in 1968 and contains data on every karyotype obtained by clinical departments performing chromosomal analyses in Denmark. The project was approved by the scientific ethic committees (Videnskabsetiske Komitéer, Reg.nr: H-B-2009-026) and the Danish Data Protection Agency (Datatilsynet, Reg.nr: 2007-58-0015).

