## Supplementary figures and images for "Metabolic signature of the pathogenic 22q11.2 deletion identifies carriers and provides insight into systemic dysregulation"

### Supplementary Data 1

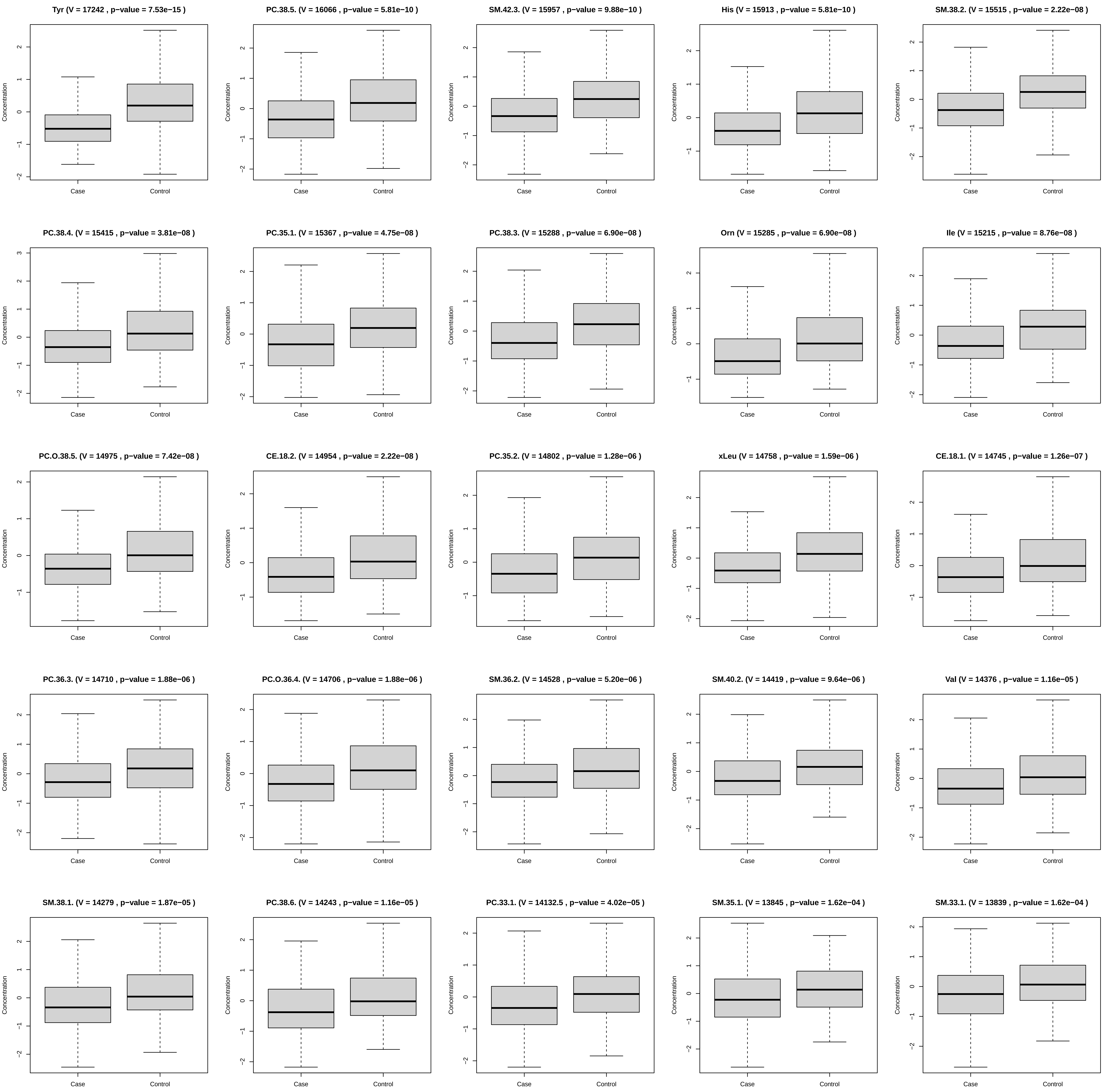

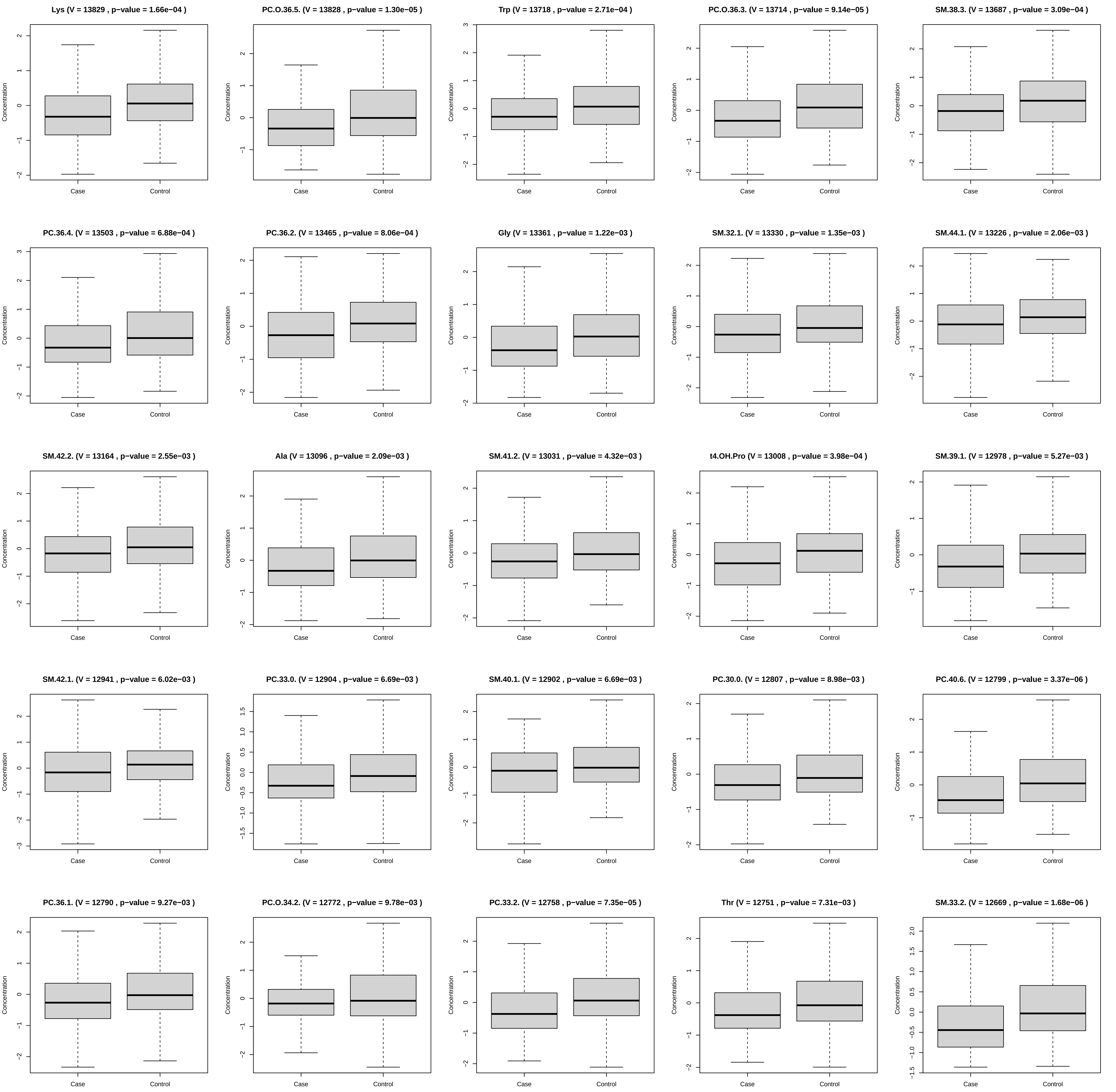

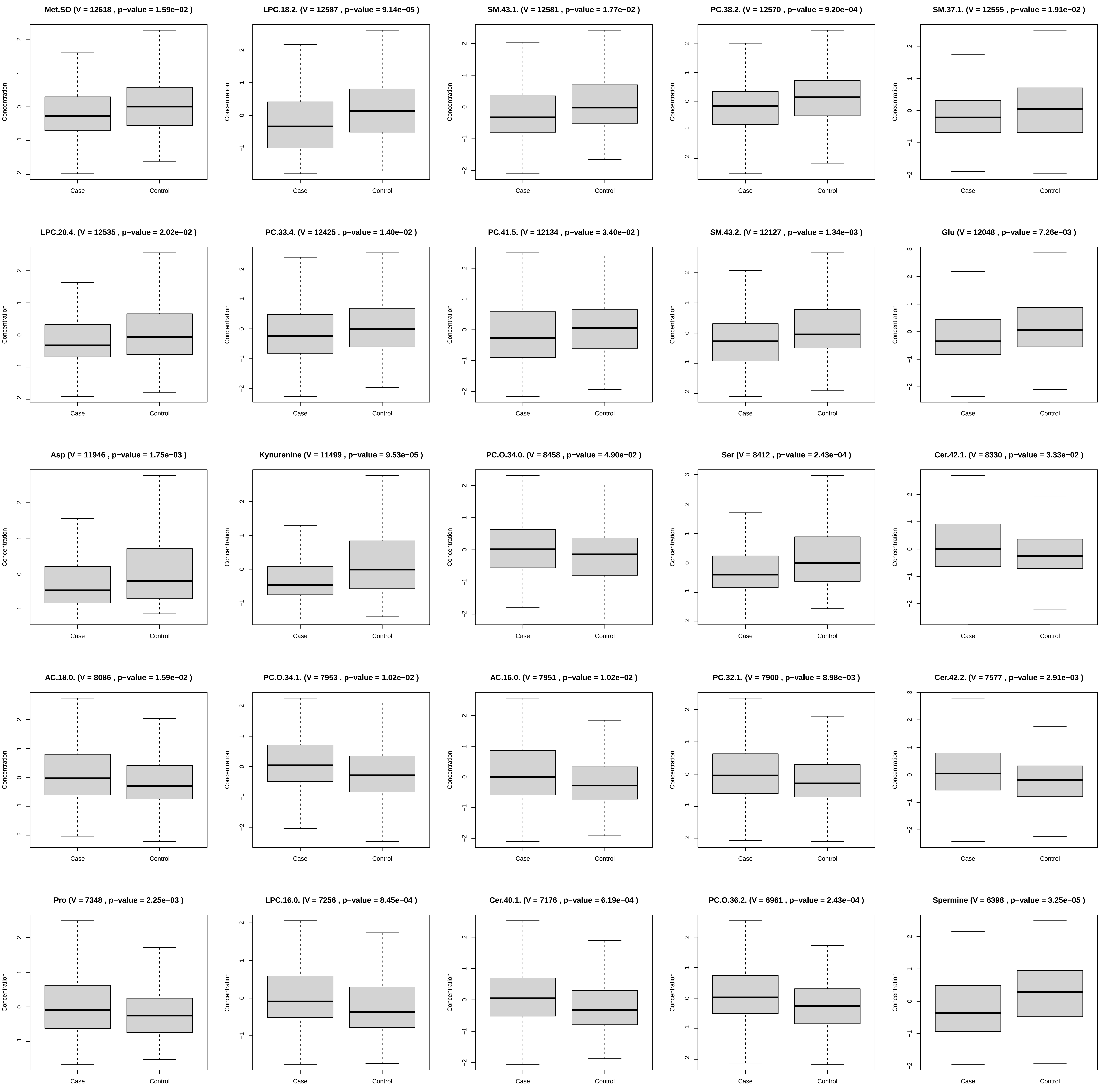

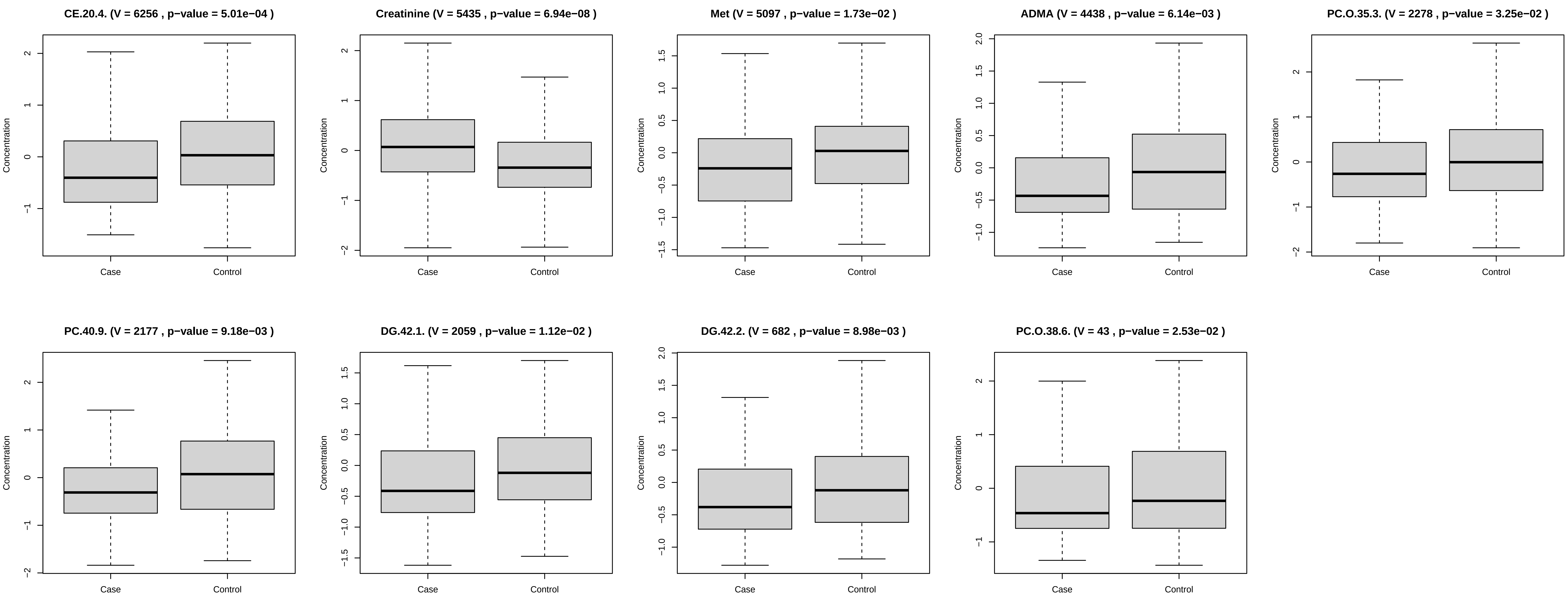
